# Aging-related matrix metallopeptidase 10 and osteopontin levels are associated with pathology, cognitive decline and age at onset in Alzheimer’s disease

**DOI:** 10.1101/2025.06.01.25328761

**Authors:** Bryan Ng, Eleftheria Kodosaki, Elena Veleva, Ashvini Keshavan, Jonathan M. Schott, Amanda J. Heslegrave, Nick C. Fox, Henrik Zetterberg

## Abstract

**INTRODUCTION:** Aging is the strongest risk factor for Alzheimer’s disease (AD) characterized by amyloid-β (Aβ) plaques and tau tangles in the brain. We aim to compare biological aging-related biomarkers among AD, non-neurodegenerative control (NDC) and non-AD neurodegenerative disease (Non-AD) individuals to evaluate their clinical utility.

**METHODS:** We included 137 participants (37 NDC, 67 AD, 33 Non-AD) from the UCL Dementia Research Centre and measured matrix metallopeptidase 10 (MMP-10), osteopontin (OPN), neurofilament-light, and glial fibrillary acidic protein in cerebrospinal fluid (CSF) samples in addition to Aβ/pTau and clinical parameters.

**RESULTS:** Elevated MMP-10 associated with poorer cognition and later onset specifically in AD, whereas elevated OPN associated with CSF Aβ and tau pathology. MMP-10 and OPN levels improved the differentiation of AD from NDC, and AD from Non-AD, respectively.

**DISCUSSION:** Our study provides evidence on potential clinical utility of CSF MMP-10 and OPN in AD diagnosis and supports taking biological aging into consideration in AD research.

## Introduction

Alzheimer’s disease (AD) is characterized by the accumulation of amyloid-β (Aβ) plaques and tau neurofibrillary tangles in the brain, with aging being the strongest risk factor for developing AD with a wide age range of dementia onset. The range of age at onset (AAO) can be as wide as 60 years even in patients with causative genetic mutations which result in high penetrance^1^. Yet, it is possible for centenarians to present with accumulations of AD pathological proteins in the brain whilst still being asymptomatic^2^. Chronological age itself is therefore a poor predictor of risk or AAO. Biological age may offer a more relevant molecular measure of aging, but it is currently unclear how biological aging modulates AD pathogenesis and progression.

Accelerated biological aging has been demonstrated to increase the risks for subsequent cognitive decline and development of dementias^3^. Cellular senescence is one of the major hallmarks of biological aging and in animal models of AD, clearing senescent glial cells in the brain can prevent cognitive deficits and pathology^4,5^. These findings laid the groundwork for a current clinical trial investigating the use of senolytics in AD patients^6^. Conversely, exogeneous toxic tau aggregates could induce cellular senescence *in vitro*^7,8^ indicating the interplay between AD pathology and biological aging.

A major research focus is on biomarker alterations both in AD patients and in presymptomatic individuals in addition to measures of the pathological proteins such as Aβ and phosphorylated tau (pTau). Characterizing these biomarker changes could help to delineate biological processes such as neuroinflammation and cellular senescence that may be associated with AD pathology and disease progression to better stratify at-risk individuals. However, there is incomplete understanding on the effects of biological aging in modulating AD using biomarkers.

In this study, we began by testing seven aging-associated biomarkers on their relationships with AD pathology and clinical manifestation by measuring these candidate biomarkers in the cerebrospinal fluid (CSF). These candidate biomarkers include neurofilament light chain (NfL) and glial fibrillary acidic protein (GFAP) which have been extensively studied due to their associations with AD pathology^9,10^ and the levels of which increase with age^11^. In addition, we quantified matrix metallopeptidase 10 (MMP-10) which is involved in extracellular matrix remodeling and inflammation moderation and has been demonstrated to be expressed/secreted by senescent cells^12,13^. It was also reported that individuals with mild cognitive impairment (MCI) exhibit higher MMP-10 CSF levels which predict their progression to AD^14^. Osteopontin (OPN; encoded by the *SPP1* gene) is another aging-related protein that is expressed/secreted by senescent cells and has previously been shown to be involved in multisystem aging^13,15^. In the disease context, OPN was first found to be highly expressed in multiple sclerosis patients and animal models^16^ as a marker of proinflammatory cytokine responsible for macrophage recruitment before it was reported that OPN CSF levels are elevated in AD and can be used to predict MCI progression to AD^17,18^. More recently, OPN was demonstrated to be the key factor that both defines microglial disease states and drives neuroinflammation in AD/tauopathy experimental models^19–21^.

We aimed to quantify the CSF levels of these candidate aging-associated biomarkers in non-neurodegenerative control individuals (NDC), non-AD neurodegenerative disease (Non-AD) and AD patients. This work focused on the relationship between these biomarkers and clinical manifestations such as pathology, cognitive outcome and AAO, and on the utility of these biomarkers in improving differentiation of AD from NDC and Non-AD.

## Methods and Materials

### Study participants, ethics and design

Individually de-identified CSF samples were collected from 2013-2022 from patients with neurodegenerative diseases at the University College London Dementia Research Centre (NRES 15/LO/1504). All participants first visited the specialist cognitive disorders service at the National Hospital for Neurology and Neurosurgery, University College London Hospitals NHS Trust, London, UK, before giving informed written consent to research sample donation at the same time of CSF sampling. The samples were selected based on known CSF Aβ1-42 and pTau-181 levels previously measured in clinical routine to define AD pathology if both conditions are met: Aβ1-42 < 650 pg/mL and pTau-181 > 57 pg/mL for the INNOTEST assay; or Aβ1-42-to-40 ratio < 0.07 and pTau-181 > 73 pg/mL for the LUMIPULSE G assay. Non-AD conditions like frontotemporal dementia (FTD) and NDC were defined based on their clinical diagnoses plus not meeting both conditions for AD pathology. One individual met these conditions but was previously diagnosed with vascular dementia hence the individual was not assigned to the AD group. The Non-AD group consists of mostly FTD variants with fewer than one-third consisting of a mixture of semantic dementia, vascular dementia, Lewy body disease and progressive supranuclear palsy cases.

The CSF samples included in this study were measured in two partially overlapping batches:

**Batch 1:** n= 24 of NDC donors and n= 38 AD donors: AD CSF biomarkers (i.e. Aβ1-42, total tau [T-Tau], pTau-181) were mostly quantified using Fujirebio’s INNOTEST assays with 7 to 9 out of the 62 samples also quantified using Fujirebio’s LUMIPULSE G assays, depending on the specific biomarker. All samples went through one freeze-thaw cycle before the measurements.

**Batch 2:** n= 31 of NDC, n= 48 AD and n= 33 Non-AD donors where 37 of the samples overlap with the first batch: AD CSF biomarkers (i.e. Aβ1-42, Aβ1-40, pTau-181 and pTau-217) were quantified using Fujirebio’s LUMIPULSE G assays. CSF T-Tau was measured using Fujirebio’s INNOTEST assay.

Batch 1 and 2 results were merged into a common dataset consisting of n= 37 of NDC, n= 67 AD and n= 33 Non-AD samples. When merging overlapping samples, we first normalized all biomarker measurements by regressing raw values from Batch 1 against Batch 2 and then prioritized Batch 1 and LUMIPULSE G readouts for duplicate measurements in both batches.

### CSF collection and processing

Local anesthesia using lignocaine was carried out before a 22-gauge atraumatic spinal needle was used to collect CSF without active withdrawal into 2 x 10 mL polypropylene containers (Sarstedt 62610018). The CSF samples were then transported to the laboratory at ambient temperature before centrifugation at 1750 g for 5 min at 4°C. The supernatant was then collected and aliquoted into polypropylene cryovials and frozen at -80°C within 2 hours of CSF collection.

### Protein quantification methods for candidate biomarkers

NfL, GFAP, MMP-10, OPN, growth differentiation factor 15 (GDF-15) and Interleukin 6 (IL-6) were measured using R-PLEX assay kits on either the MESO SECTOR or MESO QuickPlex from Meso Scale Discovery. Briefly, the CSF samples were thawed on ice and diluted in the buffer provided and incubated for 1 h with 800 rpm shaking at room temperature for plate coating, sample incubation, and antibody incubation. The plates were then read immediately afterwards in the read buffer. Soluble Klotho was measured using an ELISA kit from IBL (27998-IBL) before reading the plates on a SPECTROStar Omega microplate reader (BMG Lab Tech). The CSF dilution factors for measurements were as follows: OPN (1 in 50); GDF-15 (1 in 5); NfL, GFAP, IL-6 and Klotho (1 in 2); MMP-10 (neat). All sample groups were randomized and blinded to the researchers running the assays.

### Statistical analysis

Normality tests using Shapiro-Wilk tests indicated that the datasets do not follow a normal distribution, hence non-parametric tests were applied for all statistical analyses. For comparisons between two groups, two-tailed Mann-Whitney test was used; for multiple groups, Kruskal–Wallis test was used with Dunn’s multiple comparison and corrected with the Holm method for pairwise comparisons; Spearman’s coefficient was used for pairwise linear correlation analyses; DeLong tests were used for receiver operating characteristic (ROC) analyses comparing the area under curve (AUC) of individual biomarkers whereas Akaike information criterion (AIC) was determined for nested ROC analyses involving multiple biomarkers. A difference of more than 2 in AIC score is considered statistically significant. For the aging-associated candidate biomarkers measured in this study, outliers were removed after sex and age adjustments when any data point falls beyond two interquartile ranges (IQR) above or below the third or first quartile, respectively. NS stands for “not significant”. \**p* < 0.05, \*\**p* < 0.01, \*\*\**p* < 0.001, \*\*\*\**p* < 0.0001 for all statistical analyses. All data were represented as mean ± SEM. Finally, statistical tests and graphing were conducted mostly in R studio v2024.09.0+375 supplemented by the GraphPad Prism v10.4.0 software. R code writing was improved by generative artificial intelligence-assisted platforms.

## Results

### MMP-10 and OPN levels are elevated specifically in AD CSF

We measured all seven aging-associated candidate biomarkers (MMP-10, OPN, NfL, GFAP, GDF-15, IL-6 and Klotho) in the Batch 1 CSF samples (see *Methods*) to address whether these candidate biomarkers exhibit differential levels in AD patients compared with NDC. NfL, GFAP, MMP-10 and OPN CSF levels were significantly higher in AD samples compared with NDC (Figure 1A) whereas IL-6, GDF-15 and soluble Klotho CSF levels did not distinguish AD from NDC (Supplementary Figure 1A). We then repeated the measurements of NfL, GFAP, MMP-10 and OPN in another batch of CSF samples (Batch 2) which included samples from Non-AD patients with other neurodegenerative conditions. The differences between AD and NDC were consistent with the Batch 1 measurements, and the Batch 2 independent measurements showed notable reproducibility with those of Batch 1 (Supplementary Figure 1B and 1C). After merging the Batch 1 and 2 results for analysis from this point onwards, we observed that the NfL and GFAP levels in the CSF could not be used to differentiate AD from Non-AD individuals whereas the MMP-10 and OPN levels in the CSF were significantly higher than those in Non-AD individuals (Figure 1B). The elevated OPN levels, in particular, were highly AD-specific, whereas OPN levels were similar between NDC and Non-AD individuals.

**Figure 1:**
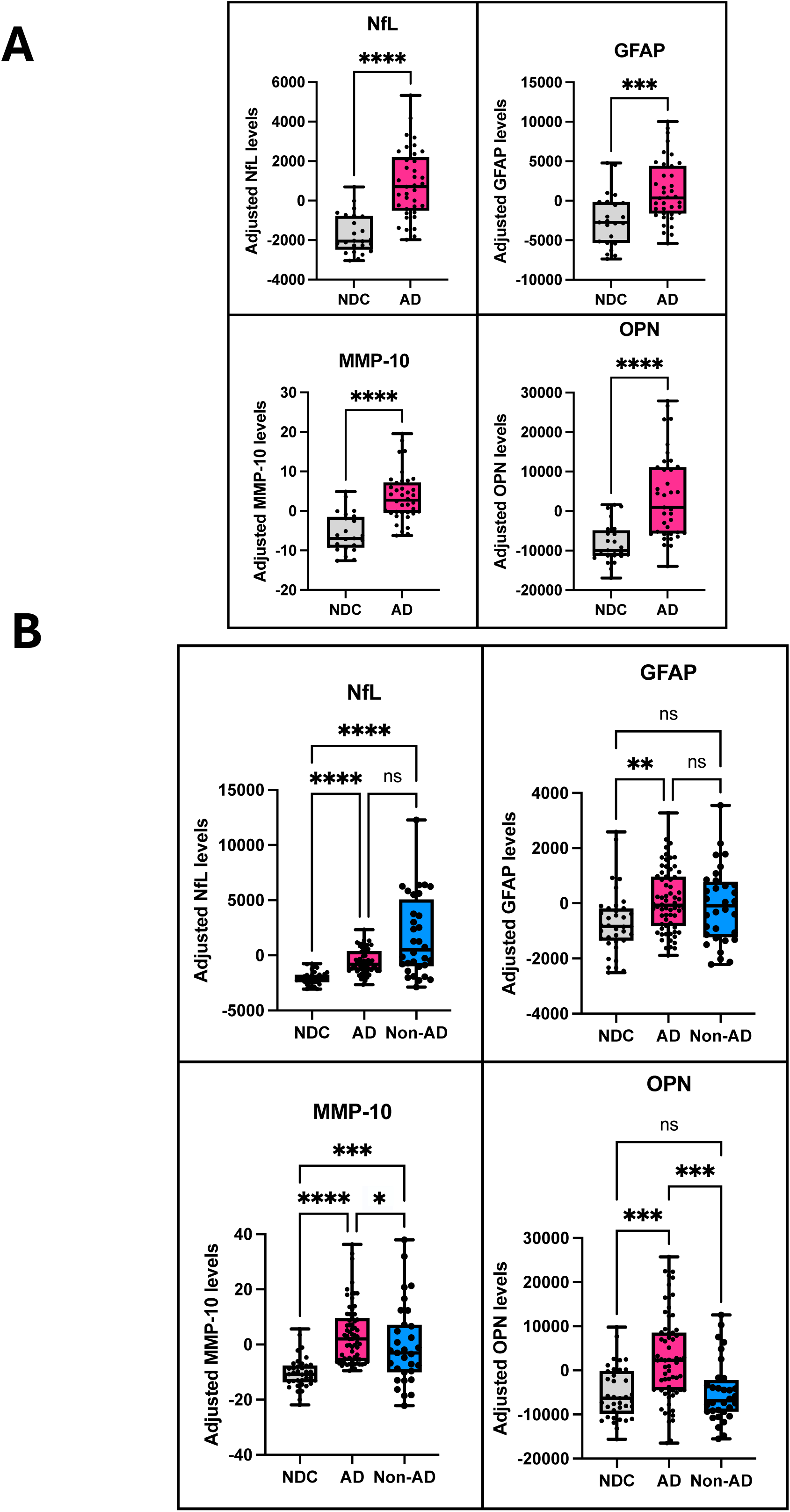
Levels of NfL, GFAP, MMP-10 and OPN in AD, NDC and Non-AD samples in the **(A)** Batch 1 CSF samples and **(B)** Batch 1 and 2 combined CSF samples. The y-axis is represented as age-and sex-adjusted levels. Boxplots indicate median, interquartile range and total range of the data. Kruskal-Wallis test was used with Dunn’s multiple comparison test.

### MMP-10 levels indicate cognitive impairment and AAO specifically in AD whereas OPN levels reflect AD pathology

We then took the results of the four candidate biomarkers forward in the context of individual cognitive impairment, age at symptom onset (AAO) and AD CSF pathology in the study population. Higher NfL levels were associated with lower MMSE scores (i.e., poorer cognitive outcomes) in AD but not in NDC, while there was a non-significant trend in Non-AD (Figure 2A). AD patients with higher MMP-10 levels also performed worse in MMSE but this relationship was not seen in NDC and Non-AD individuals. On the other hand, MMP-10 levels were linked to greater AAO only in AD while GFAP levels exhibited the same associations in both AD and Non-AD (Figure 2B). These results suggest that elevated MMP-10 levels are AD-specific (at least for the conditions included in this study) in indicating cognitive impairment and AAO as opposed to NfL and GFAP which are also elevated and associated with clinical parameters in Non-AD.

**Figure 2:**
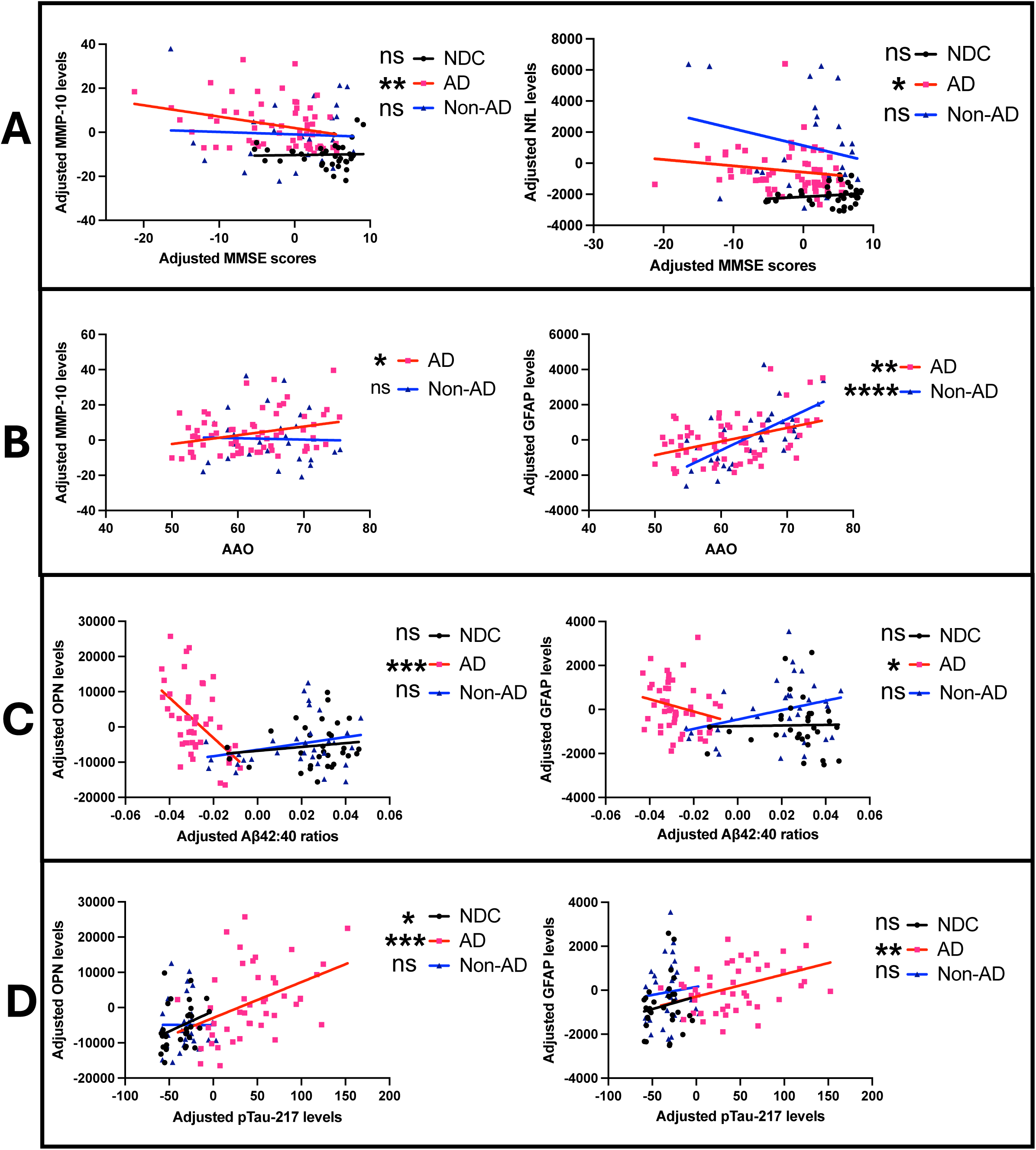
Linear correlation between CSF levels of NfL, GFAP, MMP-10 and OPN in AD, NDC and Non-AD samples in Batch 1 and 2 combined with **(A)** MMSE, **(B)** AAO, **(C)** CSF A**β**42:40 ratio and **(D)** CSF pTau-217. All axes are represented as age-and sex-adjusted levels except in **(B)** where the Y-axis values are adjusted for sex only for comparison with AAO. Spearman’s test for correlation was used.

MMP-10 levels, however, did not correlate with the AD CSF biomarkers tested in this study in both NDC and AD. This contrasted with OPN levels which were strongly linked to AD CSF pathology. Both OPN and GFAP levels were inversely correlated with Aβ42:40 ratio suggesting that greater OPN and greater GFAP levels reflect worse pathological Aβ burden specifically in AD (Figure 2C). This is especially so for the OPN levels with a more pronounced correlation with pathological Aβ burden. In terms of tau pathology in response to amyloid pathology, although the OPN levels were associated with higher pTau-217 levels in both NDC and AD, this relationship was absent in Non-AD (Figure 2D). This contrasts with the GFAP levels which correlated with pTau-217 levels in an AD-specific manner. The full correlation matrices between the four candidate biomarkers and clinical parameters/AD CSF biomarkers can be found in Supplementary Figure 2.

### Inclusion of MMP-10 improves discrimination of AD from NDC while that of OPN differentiates Non-AD patients

Finally, we asked whether CSF MMP-10 and OPN levels can improve discrimination of AD patients from NDC and Non-AD individuals. None of the four candidate biomarkers performed statistically better against the next-best biomarker or MMSE individually in differentiating AD from NDC or Non-AD (Figure 3A). MMP-10 and NfL levels individually were significantly better than OPN (p = 0.002 vs MMP-10; p = 0.004 vs NfL) and GFAP (p = 0.0005 vs MMP-10; p = 0.0009 vs NfL) levels in differentiating AD from NDC while OPN levels alone were better than GFAP levels (p = 0.003) in differentiating AD from Non-AD.

**Figure 3:**
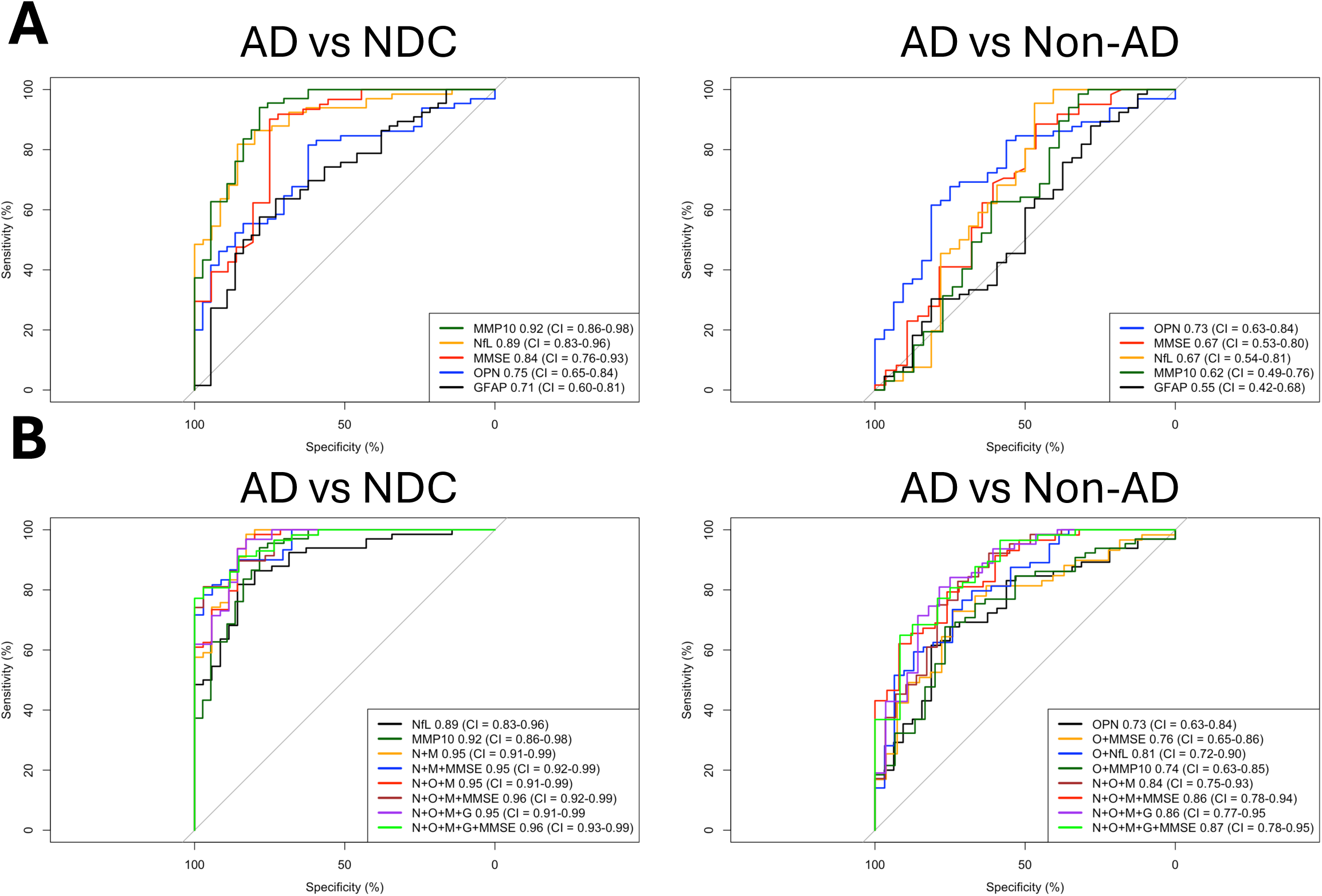
ROC analysis and corresponding AUCs from logistic regression models for the candidate CSF biomarkers and MMSE to differentiate AD patients from NDC and Non-AD either **(A)** individually or **(B)** as a nested model building upon each other using age-and sex-adjusted readouts.

To identify the most optimal combinations of these biomarkers to distinguish AD cases, we used AIC to compare between the nested ROC curves representing various combinations of biomarkers on top of the abovementioned best individual CSF biomarkers (Figure 3B). The optimal model with the lowest K value (fewest parameters) and lowest AIC score (better goodness of fit) utilized NfL, MMP-10 and MMSE together to differentiate AD from NDC (Table 2). On the other hand, including OPN into the model (i.e., NfL, MMP-10, OPN and MMSE) provided the optimal differentiating power between AD and Non-AD. This shows that apart from including NfL and MMSE as a base model, having MMP-10 alone or with OPN provide meaningful improvement in identifying AD from NDC or Non-AD, respectively.

**Table 1:** Baseline characteristics of the merged Batches 1 and 2 study population

|  | **NDC (N = 37)** | **AD (N = 67)** | **Non-AD (N = 33)** |
| --- | --- | --- | --- |
| **Demographics** |  |  |  |
| Age [Mean (SD)] | 63.8 (7.0) | 65.1 (6.5) | 67.9 (5.9) |
| Sex [Female %] | 48.6 | 56.7 | 27.3 |
| **Clinical information** |  |  |  |
| MMSE [Mean (SD)] | 26.6 (3.9) | 20.5 (6.4) | 23.9 (6.5) |
| Age at AD onset [Mean (SD)] | Not applicable | 61.9 (6.7) | Not applicable |
| **CSF AD biomarkers**<br>**(pg/mL)** |  |  |  |
| Aβ<sub>1-42</sub> [Mean (SD)(N)] | 1,280 (469) (37) | 591 (173) (65) | 970 (429) (33) |
| Aβ<sub>1-42</sub> to Aβ<sub>1-40</sub> ratio [Mean (SD)(N)] | 0.103 (0.015) (31) | 0.048 (0.01) (50) | 0.095 (0.02) (33) |
| T-Tau [Mean (SD)(N)] | 279 (111) (33) | 1,060 (427) (42) | 331 (127) (28) |
| pTau-181 [Mean (SD)(N)] | 50.5 (25.2) (37) | 179 (65.9) (66) | 44.6 (15.4) (33) |
| pTau-217 [Mean (SD)(N)] | 10.1 (7.4) (31) | 97.1 (47.5) (47) | 12.6 (9.6) (33) |
| **CSF candidate biomarkers**<br>**(pg/mL)** |  |  |  |
| NfL [Mean (SD)(N)] | 1,860 (733) (35) | 3,410 (1,050) (66) | 5,930 (3,530) (32) |
| GFAP [Mean (SD)(N)] | 2,360 (1,320) (37) | 3,320 (1,270) (66) | 3,500 (1,590) (32) |
| MMP-10 [Mean (SD)(N)] | 16.9 (5.7) (37) | 31.2 (11.1) (67) | 28.8 (14.5) (31) |
| OPN [Mean (SD)(N)] | 12,300 (6,240) (37) | 20,800 (10,400) (65) | 12,900 (6,930) (32) |

**Abbreviations:** NDC = Non-neurodegenerative controls; AD = Alzheimer's disease; Non-AD = Non-AD neurodegenerative; MMSE = Mini Mental State Examination; Aβ <sub>1-42</sub> = Amyloid- β1-42; T-Tau = Total tau; pTau = Phosphorylated tau; NfL = Neurofilament light chain; GFAP = Glial fibrillary acidic protein; MMP-10 = Matrix metalloproteinase 10; OPN = Osteopontin. This Table takes outlier removals into account. The values were rounded off to three significant figures.

**Table 2:** AIC analysis comparing nested ROC AUC parameters differentiating between AD and NDC/Non-AD groups

|  | **K** | **AIC** | **ΔAIC** | **AIC weight** |
| --- | --- | --- | --- | --- |
| **AD vs NDC** |  |  |  |  |
| **N + M + MMSE** | 4 | 54.0 | 0.00 | 0.59 |
| **N + O + M + MMSE** | 5 | 55.7 | 1.61 | 0.26 |
| **N + O + M + G + MMSE** | 6 | 57.4 | 3.33 | 0.11 |
| **N + M** | 3 | 60.4 | 6.36 | 0.02 |
| **N + O + M** | 4 | 61.6 | 7.60 | 0.01 |
| **N + O + M + G** | 5 | 63.5 | 9.45 | 0.01 |
| **AD vs Non-AD** |  |  |  |  |
| **N + O + M + G + MMSE** | 6 | 76.3 | 0.00 | 0.61 |
| **N + O + M + MMSE** | 5 | 77.3 | 1.00 | 0.37 |
| **N + O + M + G** | 5 | 84.2 | 7.87 | 0.01 |

**Abbreviations:** AIC = Akaike information criterion; ROC = Receiver operating characteristic analysis; AUC = Area under curve; AD = Alzheimer's disease; NDC = Non-neurodegenerative control; Non-AD = Non-AD neurodegenerative; N = NfL; M = MMP-10; O = OPN; G = GFAP; MMSE = Mini mental state examination. The values were rounded off to three significant figures.

## Discussion

We have demonstrated that aging-related MMP-10 and OPN in the CSF are associated with pathological and clinical manifestations specifically in AD. Our study included well-established age-associated biomarkers i.e., NfL and GFAP, to help in providing a validated reference for our findings. We confirmed that elevated MMP-10 and NfL levels are associated with worse cognitive performance as previously reported^14,22^ and further revealed that this relationship is AD-specific. We also discovered for the first time that both elevated MMP-10 and GFAP levels are linked to later AAO – in the case of MMP-10 this was AD-specific. On the other hand, OPN levels did not correlate with cognitive impairment in our study regardless of disease status, contradicting a previous report showing that CSF OPN levels increase with better cognitive performance^17^. Notably, our analysis controlled for sex and age, which both influence MMSE scores^23^, as opposed to the previous report^17^. In terms of markers of AD pathology, the marked inverse association between OPN levels and Aβ42:40 ratio we observed is consistent with a previous post-mortem report where the authors reported that OPN expression is positively correlated with Aβ burden in the brain^24^. This contrasts with MMP-10 levels, which from our results do not reflect Aβ pathology, echoing another recent study showing that MMP-10 levels do not correspond with Aβ42 levels in the CSF^25^. In addition to Aβ pathology, we demonstrated that elevated OPN levels are also tightly linked to pTau-217 levels although the relationship is also present in NDC as opposed to GFAP levels which correlate with pTau-217 levels in an AD-specific manner.

We then put these data together from a clinical perspective and included the levels of NfL, GFAP, MMP-10, OPN and MMSE scores in a binary classification model to differentiate AD from NDC or Non-AD. We demonstrated that the levels of NfL, MMP-10 and MMSE scores constitute the best predictor combination compared to NfL, MMP-10 or MMSE scores alone to distinguish AD from NDC in our study population. The addition of OPN into this combination best differentiates AD from Non-AD instead, unsurprisingly due to the strong association between OPN and Aβ pathology which is absent in the Non-AD group. Importantly, our findings show that incorporating cognitive outcomes improves the discrimination between AD and NDC/Non-AD in addition to biomarkers alone suggesting that both clinical and biological definitions of AD may work best in tandem in the clinical setting. The observations from our study have added new insights to the roles of MMP-10 and OPN in AD pathogenesis and clinical utility and substantiated the value of taking biological aging into account in the context of neurodegenerative diseases.

It has been reported previously that CSF levels of OPN increase early in AD pathogenesis and could be used to predict MCI from healthy individuals^26–28^. Taken together with the results from the current work, CSF levels of OPN appear to reflect AD pathology across disease stages. As we focused on measuring CSF samples, what we can interpret from the results is likely confined to brain aging instead of systemic aging which may be quantified using blood samples. While MMP-10 levels in the blood have not been explored in AD, plasma levels of OPN in AD are consistent with those in the CSF^29^ with one recent study demonstrating that plasma levels of OPN are associated with subsequent dementia incidence^30^. The reproducibility between investigations using CSF and plasma samples with regards to the role of OPN in AD remains to be determined.

Two biomarkers by no means define brain aging *in toto*, and future investigations into aging-related biomarkers in the context of neurodegenerative diseases can employ measurement platforms with a higher throughput to characterize all aging-related biomarkers in parallel. Nevertheless, it is worth noting that a single aging-related biomarker from CSF can improve AD identification against established biomarkers like NfL and GFAP which are more applicable to multiple neurodegenerative diseases. It is then possible that relatively focused panels may suffice to capture either brain or systemic aging using CSF or blood samples, respectively, as recently reported in an analogous study delineating the roles of five pro-youthful and pro-aging CSF factors in AD and FTD^31^. Another key consideration relates to the fact that AD patients often present with co-pathologies such as Lewy bodies that are characteristic of other neurodegenerative diseases^32^. It is critical to understand how aging-related biomarkers, such as MMP-10 and OPN, perform with single or co-pathologies as opposed to categorizing the samples into a single clinical diagnosis. Overall, the utility of quantifying aging-associated biomarkers for AD prediction, diagnosis or progression may be fully realized when these outstanding questions on disease stage, sample types, optimal biomarker combination and pathology specificity are addressed.

The significance of our work extends beyond clinical utility as the identities of the aging-related biomarkers provide clues to the cellular mechanisms which result in the changes in expression levels in the CSF. Our results highlight the specific aspect of neuroinflammation that distinguishes AD from NDC/Non-AD as both MMP-10 and OPN are expressed in the brain primarily by glial cells^33–35^ and are involved in the regulation of inflammation^12^. Putting this together with the fact that both MMP-10 and OPN are expressed by senescent cells^13^, and that senescent cell clearance mitigates AD pathology in mouse models^4,5^, elevated MMP-10 and OPN levels pinpoint a critical role of immune ageing from glial cells in AD pathogenesis and progression.

These results suggest that there may be therapeutic value in modulating MMP-10 and OPN levels. Previous reports demonstrated that the expression of MMP-9, which belongs to the same MMP family as MMP-10, can be induced by Aβ and in turn results in Aβ degradation^36,37^. On the other hand, OPN levels were reported to be elevated in macrophages engaged in Aβ clearance whereas the pharmacological inhibition of OPN impairs their Aβ uptake ability^38^. We therefore reason that the increases in MMP-10 and OPN levels in the CSF may be a protective mechanism by glial cells in an AD pathological environment, i.e., enhancing their expression levels may be a viable therapeutic strategy to mitigate AD pathogenesis.

Finally, it is important to note that there are a couple of limitations to our study. This is a cross-sectional, single cohort study which captures a snapshot of various patients who visited our memory clinic. Although we did not extend the measurements to a validation cohort, the technical validations of the measurements from overlapping samples within the same cohort were robust even when they were performed independently by two researchers. We could conclude with a high confidence level the implications of aging-related MMP-10 and OPN in symptomatic AD, but our study was not designed to assess these biomarkers longitudinally across a chronological age range of the same participants. Doing this would strengthen the study to further elucidate the interaction between aging-related biomarkers and incipient AD onset.

Our study population was selected based on both their CSF pathology and clinical diagnosis spread between fifty and eighty years of age to accurately distinguish AD from Non-AD samples across ages, but this inevitably results in a bias where each clinical diagnosis group consists of a relatively homogeneous population with preselected pathological status. This in return results in a higher AUC baseline for the ROC analyses presented in this work that is not comparable to other exploratory biomarker studies. Since MMP-10 and OPN significantly improve clinical “diagnosis” of AD from NDC/Non-AD when each diagnosis group was confirmed with CSF pathology *a priori*, we reason that these biomarker levels are highly specific in responding to AD-related changes. However, it remains to be determined how CSF MMP-10 and OPN perform in differentiating AD diagnosis from NDC/Non-AD in the wider clinical setting without the knowledge of individual pathology.

Overall, our work underlines the differential expression levels of CSF MMP-10 and OPN in AD and uncovers their clinical utility in reflecting cognitive impairment/AAO and AD pathology, respectively. Linking these results with the roles of MMP-10 and OPN in glial aging indicates that enhancing MMP-10 and OPN expression may potentially be beneficial in mitigating AD pathology. Our study outcomes warrant further investigation to take brain aging into consideration in AD research.

## Data Availability

Biomarker data produced in the present study are available upon reasonable request to the authors.

## Acknowledgements

BN was supported by the National Science Scholarship from the Agency for Science, Technology and Research, Singapore and the UK Dementia Research Institute. EK is supposed by the UK Dementia Research Institute. AK is supported by the Blood Biomarker Challenge grant to the ADAPT study (ARUK-BBC-2023-02; PIs Schott and Keshavan), and the National Institute for Health and Care Research University College London Hospitals Biomedical Research Centre. NF acknowledges support from the National Institute for Health and Care Research University College London Hospitals Biomedical Research Centre, the UK Dementia Research Institute at UCL (UKDRI-1003), Rostrees Trust and the Alzheimer’s Society. HZ is a Wallenberg Scholar and a Distinguished Professor at the Swedish Research Council supported by grants from the Swedish Research Council (#2023-00356, #2022-01018 and #2019-02397), the European Union’s Horizon Europe research and innovation programme under grant agreement No 101053962, Swedish State Support for Clinical Research (#ALFGBG-71320), the Alzheimer Drug Discovery Foundation (ADDF), USA (#201809-2016862), the AD Strategic Fund and the Alzheimer’s Association (#ADSF-21-831376-C, #ADSF-21-831381-C, #ADSF-21-831377-C, and #ADSF-24-1284328-C), the European Partnership on Metrology, co-financed from the European Union’s Horizon Europe Research and Innovation Programme and by the Participating States (NEuroBioStand, #22HLT07), the Bluefield Project, Cure Alzheimer’s Fund, the Olav Thon Foundation, the Erling-Persson Family Foundation, Familjen Rönströms Stiftelse, Stiftelsen för Gamla Tjänarinnor, Hjärnfonden, Sweden (#FO2022-0270), the European Union’s Horizon 2020 research and innovation programme under the Marie Skłodowska-Curie grant agreement No 860197 (MIRIADE), the European Union Joint Programme – Neurodegenerative Disease Research (JPND2021-00694), the National Institute for Health and Care Research University College London Hospitals Biomedical Research Centre, the UK Dementia Research Institute at UCL (UKDRI-1003), and an anonymous donor.

---

**Supplementary Figure 1:**
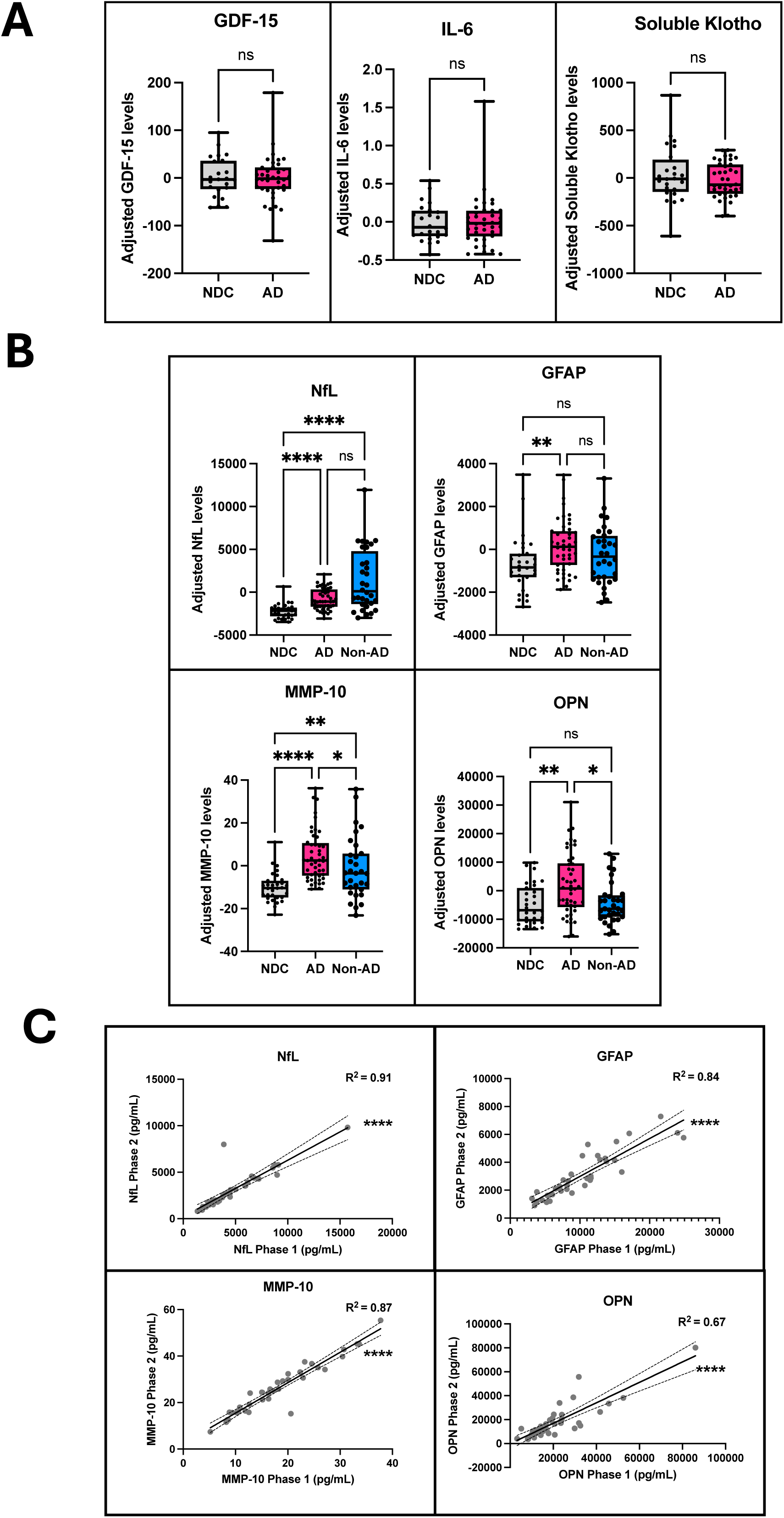
**(A)** Levels of GDF-15, IL-6 and Soluble Klotho in AD and NDC in the Batch 1 CSF samples and **(B)** Levels of NfL, GFAP, MMP-10 and OPN in AD, NDC and Non-AD in the Batch 2 CSF samples. For **(A-B)**, y-axis is represented as age-and sex-adjusted levels. Boxplots indicate median, interquartile range and total range of the data. Kruskal-Wallis test was used with Dunn’s multiple comparison test. **(C)** Linear correlation of NfL, GFAP, MMP-10 and OPN levels between Batch 1 and 2 samples. Spearman’s correlation test was used.

**Supplementary Figure 2:**
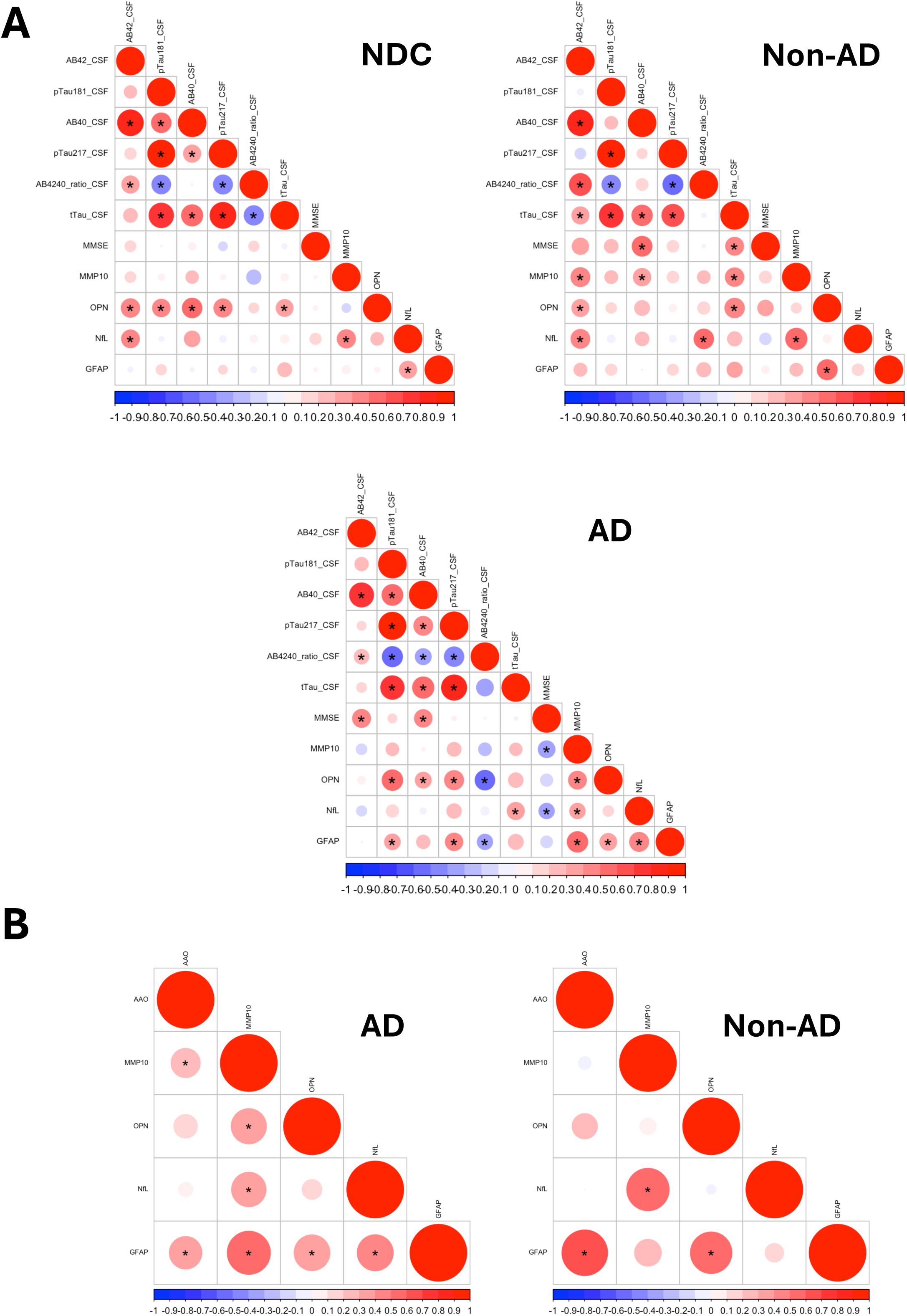
Full correlation matrices within NDC, AD and Non-AD samples between all pairwise parameters when the dataset either controlled for **(A)** both age and sex, or **(B)** controlled for sex only specifically for assessing AAO. Spearman’s correlation test was used. Asterisks indicate statistical significance; color shade indicates the value of correlation coefficient; size indicates the linear regression slope.

